# GERBEHRT: A BERT-based Model Tailored for German Electronic Health Records – Potential in Chronic Kidney Disease Prediction

**DOI:** 10.1101/2025.10.24.25338721

**Authors:** Anja Seidel, Edgar Steiger, Friedrich A. von Samson-Himmelstjerna, Lars E. Kroll

## Abstract

Routinely collected electronic health records (EHRs) contain rich longitudinal information that enables the prediction of patient outcomes at scale. We developed GERBEHRT, a transformer model adapted from BEHRT and specifically tailored to German EHRs. GERBEHRT was pretrained on outpatient claims from more than 9 million statutorily insured patients and fine-tuned with nearly 1 million additional patients to predict chronic kidney disease (CKD) - a serious condition whose progression can be delayed by early detection. GERBEHRT incorporates EHR features not previously explored in BERT-based approaches and introduces an efficient method to represent multiple attributes per medical concept, such as diagnoses and medications. In a test cohort of 3.7 million patients with 1.5% CKD positives, GERBEHRT achieved an area under the receiver operating characteristic curve (AUROC) of 87.9 and an average precision (AVPR) of 11.4 for a three-year prediction of incident moderate-to-severe CKD, outperforming riskfactor-based models (AUROC/AVPR: 83.6/6.4) and more traditional algorithms using the full EHR (AUROC/AVPR: 86.9/10.1). Although CKD risk prediction remains challenging, GERBEHRT’s superior performance underscores the importance of comprehensive EHR utilization and highlights the potential of tailored deep learning models for personalized CKD risk prediction and targeted patient screening.

## I. Introduction

**C**HRONIC KIDNEY DISEASE (CKD) is a major public health problem [1]. The disease is defined as “abnormalities of kidney structure or function, present for a minimum of 3 months, with implications for health” [2] and divided into five diagnostic stages. In 2017, the global prevalence of CKD was estimated to be 9.1% (all stages) [3] and the disease was ranked as the 12th leading cause of death [3]. In Germany, 2.3% of adults aged 18 to 79 years were estimated to be affected by CKD stage 3 or higher [4] and in 2014, 10.2% of the statutory German health insurance expenditures were estimated to be spent for patients with CKD stage 3 or 4 and 1.6% for dialysis patients [5]. Although the first stages of CKD are largely asymptomatic, medical symptoms gradually increase until dialysis or kidney transplantation may become necessary to prevent death [2]. The relatively late onset of symptoms makes early detection of CKD difficult. However, early detection combined with the reduction of risk factors through lifestyle changes or medical treatment could be an important step to slow the progression of the disease [6], [7]. The global organization “Kidney Disease: Improving Global Outcomes” (KDIGO), which develops evidence-based clinical practice guidelines in kidney disease, recommends population screening for CKD based on specific risk factors [8]. Although useful and easily applicable, there is potential in two regards. First, large-scale electronic health records (EHRs) are collected routinely in Germany and many other countries. Although not created initially for medical research but for billing purposes, they provide valuable information on the patient’s medical history and could be used in their entirety to predict the risk of developing certain diseases such as CKD. In addition to their higher information content, they remove the dependency of having to rely on manually curated risk factors, the quality of which can vary greatly depending on the clinical picture examined. Secondly, with advances in machine learning, there are ever better ways of using these information-rich but also complex EHRs to uncover hidden disease patterns and using them for increasingly accurate predictions. This raises the question of how well CKD prediction based on largescale EHRs using algorithms specially tailored for this type of data is possible and, by choosing a study design that mirrors reality as closely as possible, what the potential practical use would be. The latter could be, for example, the support of physicians in identifying patients with high risk of CKD. The electronic patient record (ePA) [9], [10], which gives people with statutory health insurance in Germany digital access to their complete health data, opens up new opportunities for risk communication between doctors and patients.

Reliable prediction of CKD has already been the objective of numerous studies. However, many rely on predefined risk factors as predictors (e.g. [11]–[15]) or use laboratory values and/or vital signs (e.g. [13], [16]–[21]) that are not collected routinely for all patients. The prediction of incident CKD based on large-scale EHR, using the entire set of features that is collected routinely, such as demographics, diagnosed diseases, and prescribed drugs, has only been tackled by few studies [22]–[25]. However, these studies chose an artificial setting in which CKD patients were overrepresented in their test data [22]–[25] compared to real CKD frequencies and/or applied age and reported sex propensity score matching [23]–[25]. This makes it difficult to assess the practical use of these models in a real-world setting, where the majority of patients are CKD negative [3] or have undiagnosed CKD [26]. Recently, CKD and other diseases were predicted for the next visit or subsequent diagnosis of a patient based on historical diagnoses by large language models (LLMs) [27]. LLMs were shown to slightly outperform Med-BERT [28], one of the first proposed “Bidirectional Encoder Representations from Transformers” (BERT) [29]-based architectures for EHR (increase of AVPR for CKD of 0.62%). It will be interesting to see whether this performance gain persists when predictions are made over a longer period of time, which would be more relevant from a patient perspective, and how LLMs scale compared to BERT-based models when additional information is considered, such as prescribed medications.

In this study, however, we want to focus on the further development of BERT-based approaches. BERT originally became famous for its excellent performance in natural language processing (NLP) tasks, but soon the first adaptions specifically developed for EHRs were published, such as G-BERT [30], BEHRT [31] or Med-BERT [28]. Due to their promising performance gains over more traditional approaches, they have been further improved over the years, for instance, by adapting the pre-training and fine-tuning objectives, the architecture, or the input features used (e.g. [32]–[39]). In our study, we had access to outpatient claims of diagnosed diseases and prescribed medications from millions of people with statutory health insurance in Germany. Although numerous BERT-based models have been developed using electronic health records in countries such as the United states (US, e.g. [28], [33], [34], [36], [37]), many healthcare systems, including Germany, do not yet have comparable, customized models. Although Germany and the US are high-income and industrialized countries, they differ from each other in many aspects of their healthcare systems and population characteristics [40]–[42]. Examples include, to name just a few, the main insurance system used (US: private health insurance, Germany: statutory health insurance) [41], access to healthcare (e.g. affordability) [40], healthcare outcomes (e.g. mortality of certain diseases [40], life expectancy [41]), the burden of diseases (e.g. obesity [41]) or demographics (e.g. rate of population aging [42]). In addition, although the disease coding systems of most countries are based on the “International Classification of Diseases” (ICD) by the World Health Organization (WHO) [43], there are country-specific variants with modified and extended diagnosis codes and additional coding properties, such as the “ICD German Modification” (ICD-GM) [44]. All of this indicates that optimal prediction performance requires models that are explicitly tailored to the structure, coding practices, and clinical characteristics of each nation’s EHRs, rather than relying on models trained elsewhere. Therefore, in this study, we developed a machine learning model designed precisely for this purpose, using Germany as a case study for nation-specific EHR modelling. We named our model “GERBEHRT” (pronounced “Gerbert”, a Germanic given name) to indicate that it was trained on German (GER) EHRs and that it extends BEHRT [31]. We incorporated input features that, to the best of our knowledge, had never been tested before, namely the specialty group of the physician, the medical substance, and the certainty of diagnosis (a special coding characteristic in Germany that indicates how confident the physician was when making the diagnosis). While previous proposed BERT-based methods characterize diseases and drugs with a single code, we developed a new strategy that allows a more powerful description of each of these concepts by various features. We assessed the final importance of these features using an ablation study. GERBEHRT was pre-trained from scratch in a semi-supervised way using longitudinal outpatient claims from more than 9 million patients with statutory health insurance in Germany. In this pre-training phase, basic patterns of diseases and medications are learned. The resulting GERBEHRT model was further fine-tuned for the prediction of moderate-to-severe CKD in the next three years using claims from nearly 1 million patients. We compare the predictive power of GERBEHRT with those of more traditional algorithms using EHRs or established risk factors. By using real-world data of millions of patients and by choosing a study design that reflects reality as closely as possible, our study allows a reliable assessment of the predictive power of the algorithms examined and thus a realistic evaluation of potential applications and their challenges in practice.

Our paper is structured as follows: first, we describe the used EHRs, then our proposed GERBEHRT architecture, and finally the CKD prediction based on risk factors, the whole EHR and GERBEHRT.

## II. Methodology

### A. Electronic Health Records

#### 1) Data Source and Pre-Processing

In Germany, health insurance is mandatory and about 87% of the population is covered by Statutory Health Insurance (SHI) [45]. In this study, we used SHI claims data for ambulatory care in Germany, specifically drug prescription data from pharmacies and other institutions according to §300 paragraph 2 German Social Code Book V, as well as diagnosis data of SHI-accredited physicians or psychotherapists according to §295 paragraph 2 German Social Code Book V. Patients were already tagged with quality labels that indicated whether they could be unambiguously assigned to the drug prescription and diagnosis data set and whether their transmitted age and reported sex were plausible. For all subsequent tasks, we considered only patients who met these plausible criteria. The two data sets do not allow for an unambiguous mapping of the diagnosed diseases and (potentially) prescribed drugs at a patient’s medical visit. However, both data sets provide the quarter (highest common time resolution) and the specialty group of the physician. Note that this limited temporal resolution means that diseases and medications within a quarter are order-invariant and should be treated as such by subsequent models.

#### 2) Features

Per patient, the diagnosed diseases were described by their 4-digit code from the German Modification of the 10th revision of the International Classification of Diseases (ICD-10-GM, n=9.098) [44] and their diagnostic certainty (n=2). The diagnostic certainty is a special coding property in Germany that is used in the ambulatory sector. We considered the commonly used attributes “confirmed” and “suspected”. The latter means that the diagnosis cannot yet be regarded as confirmed or excluded according to current medical-scientific guidelines. The prescribed medications were represented by their 3-digit code from the Anatomical Therapeutic Chemical Classification System (ATC, n=100) [46] and their main medical substance (n=2.589), derived from the pharmacy central number, a unique drug identifier used in Germany. This combination was chosen to provide information on which substance was prescribed for which reason (ATC code). Note that the same substance can be used in different doses and dosage forms to treat very different diseases and can therefore be assigned to different ATC codes. Additional features used are the patient’s age (annual resolution available) and the quarter (value between 1 and 4) at the time the disease/medication was coded, as well as the specialty group of the physician (n=18) who diagnosed the disease/prescribed the drug. While age is a known risk factor for various diseases and the quarter encodes seasonal information, combining the two can increase the temporal resolution of the recorded diagnoses and substances, as age alone is only available in annual resolution.

### B. GERBEHRT

In this section, we introduce our proposed architecture named GERBEHRT, describe how it is tailored for (German) EHR and how it can be pre-trained for subsequent tasks such as the prediction of specific diseases.

#### 1) Architecture

GERBEHRT is an extension of BEHRT [31]. The input of our model consists of two types of concepts: diseases and medications, each can be characterized by different features. Some features are shared between concept types: In our work, these are the patient’s age at the time the concept was coded, the quarter, and the specialty group of the physician who diagnosed the disease/prescribed the drug. Other features are concept-type specific: We characterize a disease by its 4-digit ICD-10-GM code and diagnostic certainty. A medication was described as a combination of its main substance and its three-digit ATC code.

The creation of the input embedding that is fed into GERBEHRT is illustrated in Fig. 1. Per patient, diagnosed diseases and prescribed drugs were ordered first by time via the patient’s age (annual resolution) and quarter (1 to 4) and then by the specialty group of physicians. In contrast to previous work (for example, [28], [31]), it is unknown which codes belong to which medical visit. Therefore, our positional embedding distinguishes between events defined as the collection of patient’s diagnoses and prescriptions for a specific age, quarter, and specialty group of the physician and treats all codes within an event as order-invariant. Similarly to BERT, per input position i, the input representation 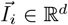, with d=hidden size, is calculated as a sum of the embeddings of the relevant input features (each ∈ ℝ^*d*^) at that position, more precisely:

**Fig. 1.**
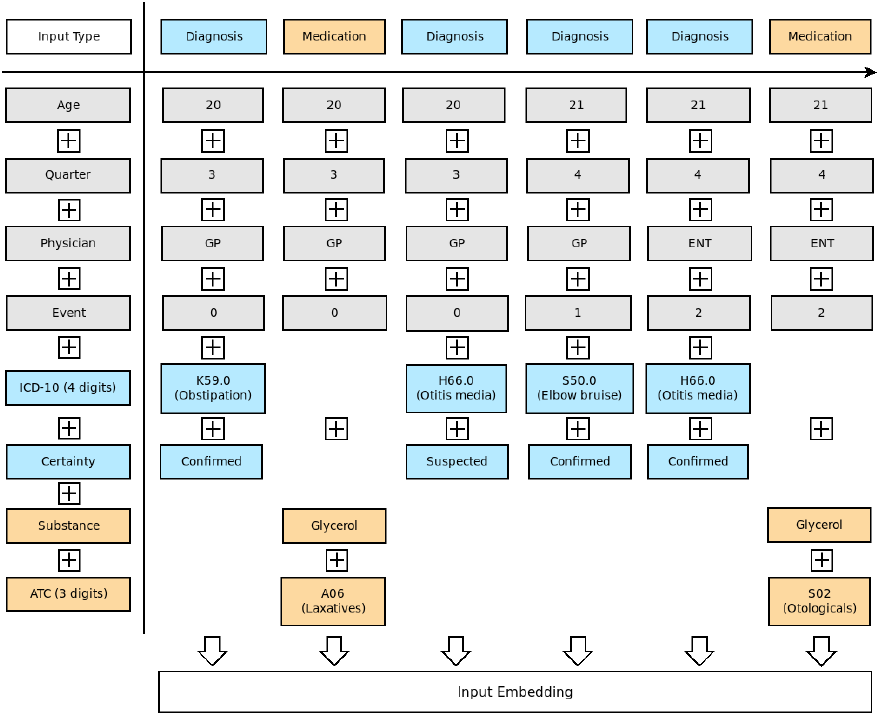
Generation of a patient’s input embedding used by GERBEHRT. Abbreviations: GP=General Practitioner, ENT=Ear, Nose & Throat Doctor

##### Algorithm 1

Calculation of GERBEHRT’s input embedding

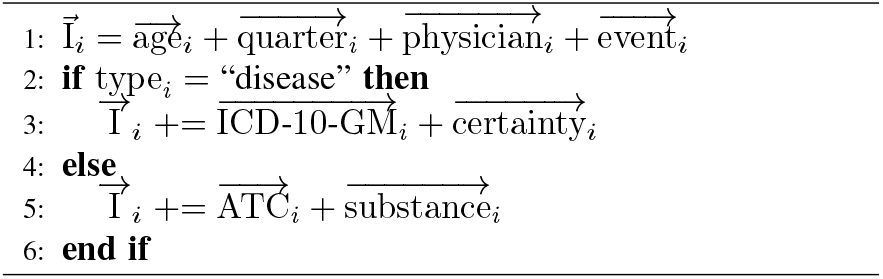

This formula allows for the description of different concept types via multiple different features. To the best of our knowledge, this is the first time a BERT-based architecture has used this strategy. The first approaches have characterized a concept only with a single code [28], [31] and recent work combined different features per concept horizontally [38] instead of stacking them vertically.

#### 2) Pre-Training and Cohort Selection

Analogous to the original BERT [29], GERBEHRT was pre-trained using masked language modeling (MLM). Due to limited computational resources, we chose the hyperparameters from BEHRT (number hidden layers, number attention heads, etc.) which Li et al. [31] determined by an extensive hyperparameter search. For the MLM task, 9.5 million patients with at least five doctor visits between 2016 and 2020, determined from the diagnostic data set, were randomly sampled. After combining diagnoses and drug prescription data per patient, quarter, and physician’s specialty group, we excluded patients with conflicting reported sexes from the two sources and patients with less than five diagnoses and ATC codes combined (which can be caused by diagnoses without the label “confirmed” or “suspected”). The characteristics of the remaining 9.485.112 patients with respect to the input features used by GERBEHRT are shown in Table I. GERBEHRT follows the approach originally proposed by BERT [29] and already applied by BEHRT [31], namely random masking of 15% of ICD-10-GM codes (since our final goal is the prediction of diseases). However, as previously described, the disease codes per patient are order-invariant within an event (the same age, quarter, and specialty group). For that reason, masking of multiple disease codes per event could lead to an erroneous loss value if the model predicts the correct codes at a wrong position within an event. To avoid this problem, we allowed for the masking of at most one disease per event.

**TABLE I.** Characteristics of the population used for Masked Language Modeling (9.485.112 PATIENTS, MALE: 46.9%, FEMALE: 53.1%). Per patient, we computed properties of different features from the patient’s EHR between 2016 and 2020 (with n=number/count) and built the mean, median and standard deviation (SD) over the entire population.

| Feature | Property | Median | Mean (SD) |
| --- | --- | --- | --- |
| Age | At first medical event | 44 | 42.38 (24.38) |
| Quarters | n(with medical events) | 16 | 14.75 (5.25) |
| Diagnoses | n(total) | 89 | 151.42 (173.07) |
|  | n(confirmed) | 85 | 147.20 (170.92) |
|  | n(suspected) | 2 | 4.22 (6.11) |
|  | n(distinct ICD-10-GM<sup>a</sup>, level 4) | 32 | 37.79 (24.76) |
| Drugs | n(total) | 19 | 37.00 (46.09) |
|  | n(distinct ATC<sup>b</sup>, level 3) | 6 | 7.11 (4.90) |
|  | n(distinct substances) | 8 | 9.70 (7.85) |
| Physicians | n(distinct groups) | 5 | 5.42 (2.50) |

<sup>a</sup> German Modification of the 10th Revision of the International Classification of Diseases, version 2021 [44]
<sup>b</sup> Anatomical Therapeutic Chemical Classification System [46]

Our models were trained on a single NVIDIA GeForce RTX 2080 Ti with a batch size of 32. The publicly available BEHRT source code [31] served as the basis of our implementation. Since BEHRT was built using the now obsolete Huggingface pytorch-pretrained-bert library, we migrated to the current Huggingface transformer library [47].

#### 3) Clustering of Disease Codes

Similarly to BEHRT, we visualized the disease embeddings learned by GERBEHRT, specifically the weights of the first hidden layer of the ICD-10-GM codes. For this, we reduced the dimension of the 288-dimensional ICD-10-GM codes learned by GERBEHRT with UMAP, clustered the lower-dimensional ICD-10-GM codes with HDBSCAN [48], visualized the clustering with a twodimensional UMAP projection, and labeled clusters in which at least 50% of ICD-10-GM codes belong to one ICD chapter with that chapter. We used UMAP to reduce the dimension since HDBSCAN is a density-based clustering algorithm that performs poorly in high dimensions due to sparsity. To assess the quality of the clustering, we computed the V-Measure [49] based on the ICD chapters of the clustered codes. As ICD-10-GM chapters XVIII, XX, XXI and XXII do not contain concrete thematic areas, the ICD-10-GM codes of these chapters (n=868) were excluded from the cluster evaluation (but not from the subsequent visualization). The best hyperparameters were determined with a grid search over the UMAP parameters n components (min: 2, max: 10, step size: 2), n neighbors (5, 30, 5), min dist (0, 0.1, 0.05) and metric [“euclidean”, “cosine”] and HDBSCAN parameters min samples (5, 20, 5) and min cluster size (5, 45, 5), on condition that at least 75% of ICD-10-GM codes (ignoring chapter XVIII, XX, XXI and XXII) should be clustered.

#### 4) Ablation Study

To assess the importance of GERBEHRT’s input features, we excluded a single feature at a time by removing the corresponding embedding layer, retrained GERBEHRT via MLM from scratch and compared the performance of the resulting model with the one with all features included on the CKD prediction task, which is described in the next section.

### C. Prediction of Chronic Kidney Disease

We conducted a retrospective study and predicted the risk of developing moderate-to-severe CKD. In the following sections, we describe our study design, the risk factors used as predictors, as well as how predictions were performed using the whole EHR with GERBEHRT and more traditional machine learning algorithms, respectively.

#### 1) Study Design and Cohort Selection

We predict whether a patient was first diagnosed with moderate-to-severe CKD (stage 3 or higher) between 2018 and 2020, using an observation period of three years between 2014 and 2016, and one year between as a “data gap” year (in analogy to [23]) to simulate the delay in time before claims data are available for prediction. We considered only patients of at least 35 years of age at the prediction time point (“data gap” year), as people with statutory health insurance from this age are entitled to a regular health check-up in Germany. All patients included for the prediction of CKD should have a presumed therapeutic benefit. Therefore, patients are excluded if they have already been diagnosed with any stage of CKD or are already being treated according to medical guidelines. Specifically, this means that none of the patients considered for prediction had a diagnosis of CKD (ICD-10-GM code: N18), unspecified kidney disease (N19), long-term dialysis (Z99.2), or a planned or performed kidney transplant (Z94.0, Z75.60, Z75.64, Z75.70, Z75.74) during the observation period and none of the patients considered had already received a combination therapy with renin–angiotensin system blockade (ATC code: C09) and sodium-glucose cotransporter 2 inhibitors (A10BK, A10BD15-16, A10BD19-21, A10BD23-25, A10BD27, A10BD29-30), as patients taking these medications are already under close medical observation and can be considered well treated [50]. To allow for prediction of the models, patients must have at least one diagnosis (confirmed or suspected) during the observation period. Patients who were already used by GERBEHRT in MLM pre-training were excluded to avoid prediction bias. This results in a population of 36.968.053 patients. Although medical claims data are subject to an intensive data cleaning and preparation process, they can contain coding and data transmission errors [51]. To reliably identify patients with incident moderate-to-severe CKD in the prediction period, we have applied the M2Q criterion [52]. This states that an outpatient diagnosis should only be considered valid if it occurs in at least two different quarters. Specifically, we have defined the following two conditions, of which at least one must be fulfilled by a patient to be considered moderate-to-severe CKD positive:

1. At least two confirmed diagnoses of moderate-to-severe CKD (N18.3-5) or “long-term dependence on dialysis for renal insufficiency” (Z99.2) in two different quarters within one year.
2. At least one confirmed diagnosis of criteria 1 and one confirmed diagnosis supporting moderate-to-severe CKD such as a request for kidney transplantation (Z75.60, Z75.64, Z75.70, Z75.74), other stages of CKD (N18.1-2, N18.8-9), unspecified kidney failure (N19) or dialysis (Z49, T82.4) in two different quarters within one year.

Patients without incident moderate-to-severe CKD were defined as those without any moderate-to-severe CKD code (N18.3-5) and without “long-term dependence on dialysis for renal insufficiency” (Z99.2), regardless of diagnostic certainty, throughout their entire history.

Using these criteria, out of our population of 36.968.053 patients, 563.733 were categorized as moderate-to-severe CKD positive and 36.323.812 as CKD negative. For 80.508 patients, it is uncertain whether they are positive or negative (e.g., a single moderate-to-severe CKD diagnosis over the entire prediction period). Therefore, the latter were excluded from the prediction task.

Patients were split into training, validation, and test data. The training data contain 80% of CKD positive patients and the validation and test data 10% each. To facilitate model training, the training data set was built in a class-balanced way (number positives=number negatives: 450.986). In contrast, the validation and test data sets were constructed in a way that represents the incidence of moderate-to-severe CKD in the next three years in our population (1.53%), resulting in heavily class-unbalanced data sets (number positives: 56.373, number negatives: 3.632.361). For details, see Table AI in the Appendix.

#### 2) Prediction Based on Risk Factors

We evaluate the prediction of moderate-to-severe CKD using common risk factors, namely age, hypertension (ICD-10-GM codes: I10-15), diabetes (E10-14), cardiovascular disease (ICD-10-GM Chapter 9, definition according to the annual US Health Report [53]), obesity (E65-E68), hypercholesterolemia (E78.0) and prior acute kidney injury/disease (AKI/AKD, N17-N19) [8], [54], [55]. For each disease-based risk factor, we check whether it occurred as a confirmed diagnosis during the observation period, resulting in six binary features. The age of the patient is considered at the beginning of the observation period. We tested three prediction models: The simplest approach checks whether any disease-based risk factor occurred during the observation period and predicts moderate-to-severe CKD in this case. In the other two approaches, prediction is made with the commonly used logistic regression (LR) and LightGBM [56] algorithms, respectively, based on the binary-coded disease-based risk factors and the patient’s age. In contrast to the “anyrisk-factor”-approach, these algorithms return estimates for the risk of developing CKD, whereby a prediction threshold can be selected to optimize for different metrics such as the F_1_ score or the Youden’s index (J) [57] (see Section “Performance Metrics”). Both algorithms were fit on the training data set using the Python scikit-learn library [58]. The hyperparameters for LightGBM, namely the number of estimators and leaves, were determined via a grid search with evaluation on the validation data set.

#### 3) Prediction Based on HER

For predictions based on the whole EHR, we applied our proposed GERBEHRT architecture (see Section “Architecture” for details about the concrete features). After pre-training via MLM (see Section “Pre-Training and Cohort Selection”), the GERBEHRT models were provided with different classification heads and finetuned on the CKD training data set. We used the average precision score (AVPR) achieved on the validation data set as the early stopping criterion. The AVPR was chosen because the validation and test data sets are heavily unbalanced. The following classification heads were tested:

- Multilayer perceptron (MLP): The pooled output of GERBEHRT (288-dimensional vector) is passed through a dropout layer (p=0.1), followed by a linear layer (200 hidden units) with ReLU activation, followed by a second linear layer (output dimension=1) with sigmoid activation.
- Convolutional neural network (CNN): The sequence of the last hidden states of GERBEHRT is processed by a 1D convolutional layer (256 filters, kernel size= 3, stride=1, padding=1) with ReLU activation, followed by a second 1D convolutional layer (128 filters, kernel size= 3, stride=1, padding=1) and global max pooling along the sequence dimension. The pooled features (128-dimensional vector) are passed through a dropout layer (p=0.3) and a linear layer (output dimension=1) with sigmoid activation.
- Long short-term memory (LSTM): The sequence of the last hidden states is fed into a bidirectional LSTM (hidden size=128). The final forward and backward hidden states are concatenated (2×128=256-dimensional vector) and passed through a dropout layer (p=0.3) and a linear layer (output dimension=1) with sigmoid activation.
- Attention: The sequence of the last hidden states are passed through a linear layer with tanh activation to compute attention scores for each token, which are normalized by applying a softmax function (resulting in a vector of size sequence length). Afterwards, a weighted sum of the hidden states is computed to form a context vector by multiplying the hidden states with the attention scores and building the sum over the sequence dimension. The 288-dimensional context vector is passed through a dropout layer (p=0.3) and a linear layer (output dimension=1) with sigmoid activation.

To evaluate the importance of the pre-training phase, we also randomly initialized a GERBEHRT model, skipped the pretraining and instead directly fine-tuned the model with the simple MLP head.

In addition, we have attempted to summarize the information from the EHR in such a way that it can be used by the more traditional approaches of LR and LightGBM. As both algorithms expect a fixed number of input features per patient, they cannot deal directly with our multi-feature sequential input data fed into GERBEHRT. Instead, per patient, we considered the age at the beginning of the observation period and binary encoded which ICD-10-GM codes (n=9.098), substances (n=2.589) and ATC codes (n=100) were recorded and which specialty groups (n=18) were visited, resulting in 11.806 input features. Note that these models lose any information regarding the coarse temporal order of codes, the information about which diseases were treated by which type of physician, the certainty of diagnosis, and the combination of substance and ATC-code.

#### 4) Performance Metrics

The models were evaluated using the following metrics:

– Area under the receiver operating curve (AUROC) [59]
– Average precision score (AVPR) [60]
– 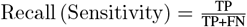
– 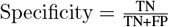
– 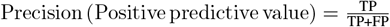
– 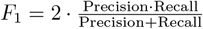
– 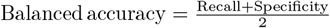

with TP=true positives, TN=true negatives, FP=false positives and FN=false negatives. As prediction cutoffs, we chose the threshold obtaining the maximum Youden’s index (J) [57], defined as J = recall+specificity − 1 and the maximum F_1_ score.

## III. Results

We have extended existing BERT-based architectures tailored for EHR in two ways. First, we have designed a new strategy that allows the characterization of different concepts (in our case, diagnoses and prescriptions) with several different features. Second, we explore features that, to the best of our knowledge, have never been tested before, namely the physician’s specialist group, medical substance, and certainty of diagnosis. Our model, called GERBEHRT, was pre-trained semi-supervised on German outpatient claims data covering 5 years from more than 9 million patients (see Table I for detailed patient characteristics). In the next section, we first visualize the disease embeddings learned by GERBEHRT, demonstrating the effectiveness of the pre-training procedure. Afterwards, we present the results that GERBEHRT achieves in a healthcare-relevant disease prediction task, namely the risk of developing moderate-to-severe CKD.

### A. Disease Embeddings of GERBEHRT

To visually verify whether our GERBEHRT model learned semantically meaningful embeddings during pre-training, we clustered its learned 4-digit ICD-10-GM code embeddings (n=9.098) and colored the resulting clusters by ICD chapter. More precisely, the original 288-dimensional ICD-10-GM code embeddings were reduced to 6 dimensions using UMAP (n neighbors=15, min dist=0, metric=“cosine”), then clustered using HDBSCAN (min samples=5, min cluster size=10) and visualized with a two-dimensional UMAP projection (see Fig. 2). A total of 71.96% of the ICD-10-GM codes could be grouped into 134 clusters. While 51.34% of the clustered codes were assigned to one large cluster, the remaining ICD-10-GM codes are distributed relatively evenly among other clusters. Clusters with at least 50% of ICD-10-GM codes belonging to the same ICD chapter and, therefore, having easily detectable semantic similarity are colored by that chapter. This applies to 107 of the 134 clusters. In addition, for 17 of the 18 ICD chapters considered, at least one cluster was found that contains ICD-10-GM codes that predominantly belong to this chapter.

**Fig. 2.**
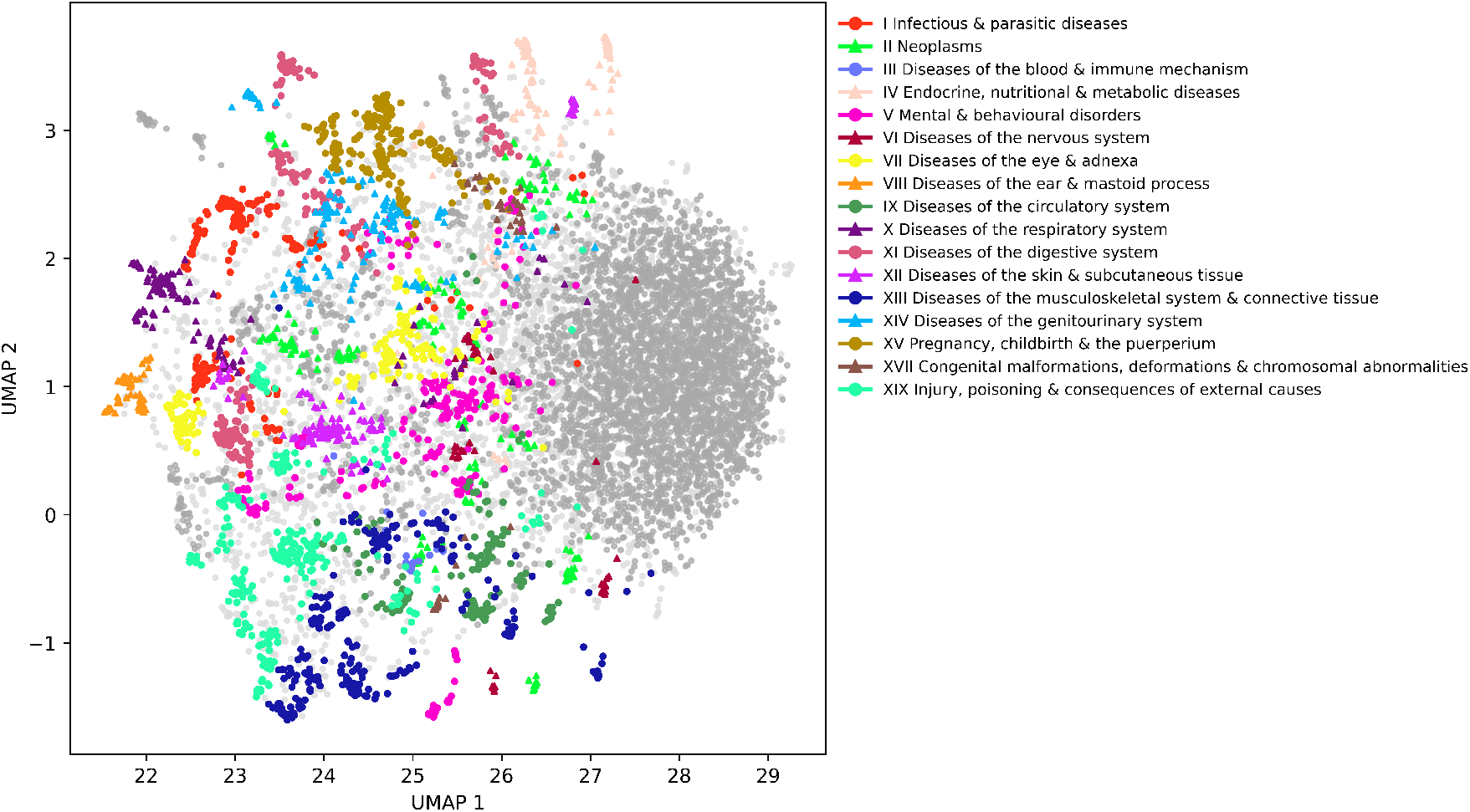
UMAP plot (n neighbors=15, min dist=0.05, metric=“cosine”) of HDBSCAN clusters of the 4-digit ICD-10-GM code embeddings of GERBEHRT. Clusters with **≥** 50% of ICD-10-GM codes belonging to a single ICD chapter are colored according to this chapter. Clusters without a dominating ICD chapter are colored in dark grey and ICD-10-GM codes that could not be clustered are marked in light grey.

### B. Prediction of Chronic Kidney Disease

We predicted whether a patient will develop CKD of stage ≥3 in the next three years, given a history of diagnosed diseases and medications prescribed over the last three years. The models were trained on data from 450.986 CKD positive and negative patients, respectively (see Table AI in the Appendix). The test data was built in a way that represents the observed three-year incidence of CKD in our population, resulting in highly unbalanced data of 56.373 (1.5%) CKD positive and 3.632.361 (98.5%) CKD negative patients.

#### 1) Predictive Power of Risk Factors

We used established risk factors, namely age, hypertension, diabetes, cardiovascular disease, obesity, hypercholesterolemia and prior acute kidney injury/disease to predict moderate-to-severe CKD. The performance metrics are shown in the section “Risk factors” of Table II. The corresponding confusion matrix counts, for example, the number of true and false positives can be found in Table AIII in the Appendix. With the simplest approach, moderate-to-severe CKD is predicted for all patients with *any* disease-based risk factor during the observation period (classifier 1). It can be shown that more than 95% (recall) of patients with moderate-to-severe CKD can be detected with this approach. However, the high recall is offset by a very low precision value of just 2.17%. This means that by simply predicting all patients with any risk factor as moderate-to-severe CKD positive, 97.83% of them would be false positives. In total, this results in a low F_1_ score (harmonic mean of precision and recall) of 4.23%. In addition to this simple approach, we made predictions based on the named risk factors (disease-based and age) using the LR (classifier 2) and LightGBM (classifier 3, n estimators: 100, n leaves: 40). In contrast to the “any-risk-factor”-approach, these models return estimates for the risk of developing CKD. While both models achieve respectable AUROC values of 83.62% (LightGBM) and 82.98% (LR), they also struggle with low precision values, resulting in low average precision scores of 6.42% (LightGBM) and 5.48% (LR). To enable a better comparison of these CKD probability-returning models with the baseline “any-risk-factor”-approach, two additional prediction thresholds were selected: one that achieves the precision and one that achieves the recall of the baseline approach. The corresponding performance values can be found in Table AII in the Appendix.

**TABLE II.** Performance metrics for moderate-to-severe CKD prediction based on established risk factors, EHR statistics aggregated over the observation period and temporal EHRS with different (machine learning) algorithms on the test data set. The highest values of AUROC and AVPR are highlighted in bold.

| Classifier | AUROC | AVPR | Cutoff | $F_1$ | Recall | Precision | Specificity | Balanced accuracy |
| --- | --- | --- | --- | --- | --- | --- | --- | --- |
| **Risk Factors** |  |  |  |  |  |  |  |  |
| 1) any (w/o age) | - | - | - | 4.23 | 95.39 | 2.17 | 33.11 | 64.25 |
| 2) LR | 82.98 | 5.48 | $F_1$ (0.77)<br>$J$ (0.48) | 10.95<br>7.96 | 39.35<br>83.11 | 6.36<br>4.18 | 91.01<br>70.45 | 65.18<br>76.78 |
| 3) LightGBM | 83.62 | 6.42 | $F_1$ (0.81)<br>$J$ (0.50) | 12.26<br>8.08 | 29.92<br>82.94 | 7.71<br>4.25 | 94.44<br>70.97 | 62.18<br>76.96 |
| **Aggregated statistics of EHRs** |  |  |  |  |  |  |  |  |
| 4) LR | 86.13 | 8.62 | $F_1$ (0.87)<br>$J$ (0.47) | 16.04<br>9.26 | 29.10<br>82.99 | 11.07<br>4.91 | 96.37<br>75.03 | 62.73<br>79.01 |
| 5) LightGBM | 86.92 | 10.05 | $F_1$ (0.86)<br>$J$ (0.51) | 17.23<br>9.62 | 28.88<br>83.20 | 12.27<br>5.11 | 96.80<br>76.00 | 62.84<br>79.60 |
| **Temporal EHR Data** |  |  |  |  |  |  |  |  |
| 6) GERBEHRT w/o pre-training + MLP | 87.51 | 10.76 | $F_1$ (0.88)<br>$J$ (0.50) | 18.05<br>9.65 | 29.63<br>84.93 | 12.98<br>5.12 | 96.92<br>75.57 | 63.27<br>80.25 |
| 7) GERBEHRT + LSTM | 87.77 | 11.15 | $F_1$ (0.91)<br>$J$ (0.48) | 18.50<br>10.01 | 27.34<br>84.36 | 13.98<br>5.32 | 97.39<br>76.71 | 62.36<br>80.53 |
| 8) GERBEHRT + MLP | 87.81 | 11.27 | $F_1$ (0.87)<br>$J$ (0.47) | 18.62<br>9.97 | 29.28<br>84.72 | 13.65<br>5.29 | 97.13<br>76.48 | 63.20<br>80.60 |
| 9) GERBEHRT + CNN | 87.90 | 11.32 | $F_1$ (0.87)<br>$J$ (0.46) | 18.62<br>9.92 | 27.52<br>84.85 | 14.07<br>5.27 | 97.39<br>76.32 | 62.45<br>80.59 |
| 10) GERBEHRT + Attention | **87.94** | **11.42** | $F_1$ (0.87)<br>$J$ (0.50) | 18.68<br>10.05 | 27.04<br>84.52 | 14.26<br>5.34 | 97.48<br>76.76 | 62.26<br>80.64 |

Abbreviations: AUROC=area under the receiver operating curve, AVPR=average precision score, $J$ =Youden's index, $F_1=F_1$ score, LR=logistic regression, MLP=multilayer perceptron, CNN=convolutional neural network, LSTM=long short-term memory, Attention=Attention layer

#### 2) Predictive Power of Electronic Health Records

We predicted moderate-to-severe CKD based on EHR statistics over the observation period, namely, which ICD-10-GM codes, substances and ATC codes were recorded and which specialty groups were visited, together with the patient’s age at the start of the observation period. The results can be found in Table II, section “Aggregated statistics of EHRs”. Again, predictions were made using the LR (classifier 4) and LightGBM (classifier 5, n estimators: 200, n leaves: 175). It can be seen that a noticeable increase in performance was achieved compared to the predictions restricted to risk factors (classifiers 1 to 3). In the case of the better performing LightGBM algorithm, the AUROC increased by 3.30% to 86.92% and the AVPR by 3.63% to 10.05%.

#### 3) Predictive Power of GERBEHRT

We developed a BERT-based model called GERBEHRT that was specially designed for German EHRs. Per patient, our models processes the information at which age, in which quarter, which group of physicians made which diagnoses with which certainty and prescribed which medication, represented by its ATC code and substance. GERBEHRT was tested with different classification heads. To estimate the value of GERBEHRT’s pre-training, a GERBEHRT model that skipped the pre-training phase was also tested. The results are shown in section “Temporal EHR Data” of Table II. It can be observed that all GERBEHRT models (classifier 6 to 10) outperform the more traditional LR and LightGBM fitted on EHRs (classifier 4 & 5), both in terms of AUROC and AVPR. However, these performance gains are rather moderate. The best performing model, GERBEHRT with the attention layer head (classifier 10), achieves an AU-ROC of 87.94% and an AVPR of 11.42%, which corresponds to an increase of 1.02% and 1.37%, respectively, compared to LightGBM (classifier 5). By choosing a prediction cutoff that optimizes the F_1_ score, GERBEHRT would detect 27.04% (recall) of CKD positives. Out of all predicted positives, 14.26% (precision) would be true positives. Using Youden’s index J instead, the recall increases to 84.52% at the cost of a low precision of 5.34%. The GERBEHRT model that was randomly initialized and fine-tuned for CKD prediction without pre-training (classifier 6) was the worst performing GERBEHRT model, which demonstrates the importance of the pre-training procedure.

#### 4) Ablation Study of GERBEHRT

GERBEHRT receives as input various features of the EHRs, some of which have been tested for the first time with a BERT-based architecture. To assess their impact on model performance, we conducted an ablation study of GERBEHRT on the CKD prediction task using the simple MLP classification head (see Table III). As expected, the GERBEHRT model that uses all features reaches the highest performance with an AUROC of 87.81 and an AVPR score of 11.27. Removing information about the patient’s age leads to the greatest performance loss (ΔAVPR: −0.55), demonstrating its high relevance to the prediction task. Excluding the quarter, substance or physician’s speciality group showed a medium impact on performance (ΔAVPR: −0.34 to −0.32). The diagnostic certainty and ATC codes were of lower importance (ΔAVPR: −0.23 to −0.20) in this prediction task. If the prediction is based exclusively on ICD codes, performance drops sharply (ΔAVPR: −1.89).

**TABLE III.** Performance of GERBEHRT with MLP classification head for CKD prediction on the test data set after excluding various input features (ordered by importance defined as loss of AVPR).

| Excluded feature | AUROC | AVPR |
| --- | --- | --- |
| - | **87.81** | **11.27** |
| Age | 87.06 (-0.75) | 10.72 (-0.55) |
| Quarter | 87.60 (-0.21) | 10.93 (-0.34) |
| Substance | 87.68 (-0.13) | 10.93 (-0.34) |
| Physician | 87.61 (-0.20) | 10.95 (-0.32) |
| Diagnostic certainty | 87.67 (-0.14) | 11.04 (-0.23) |
| ATC | 87.72 (-0.09) | 11.07 (-0.20) |
| all (except ICD-10-GM) | 86.17 (-1.64) | 9.38 (-1.89) |

## IV.DISCUSSION

### A. Context and Relevance

In this study, we investigated how reliably CKD can be predicted from large-scale EHRs using both traditional and advanced machine learning techniques. To this end, we extended the previously proposed BEHRT [31] architecture, adapting it to the structure and coding practices of national EHR data in Germany. Our model, called GERBEHRT, allows medical concepts to be represented with various different features, and in this work we assessed the value of incorporating additional types of features. We trained GERBEHRT from scratch on German outpatient claims data of diseases and medications from more than 9 million patients. The prediction of incident CKD for a practice-relevant prediction period of months or years, based on large routinely collected EHRs, including diagnoses and prescriptions but not laboratory values or clinical texts, has only been done so far by Vásquez-Morales et al. [22] and Krishnamurthy et al. [23], as well as subsequent work by Saif et al. [24], [25]. However, CKD positives are overrepresented in their test data sets compared to the real world [22]–[25], and/or age and reported sex propensity score matching was applied [23]–[25]. We, on the other hand, have tried to keep our data as close to reality as possible to provide real-world evidence and to make it easier to assess potential practical use. To this end, our validation and test data have been modeled to reflect the observed three-year incidence of CKD in our population. In addition, we considered the EHRs of a much larger number of CKD positive patients (GERBEHRT: 563.733, [22]: 20.000, [23]: 18.000), which better exploits the potential of deep learning architectures such as GERBEHRT.

### B. Findings

We found that the vast majority (>95%) of the patients, who developed moderate-to-severe CKD in the next three years, had at least one established risk factor during the observation period. However, only a fraction of patients with risk factors were actually diagnosed with moderate-to-severe CKD. Models that tried to predict CKD only based on risk factors reached high recall values, but suffered from quite low precision scores. This trend is consistent with previous work [11] and limits the use of these models in practice.

Predictions based on entire EHRs were characterized by a jump in performance compared to predictions limited to risk factors. Specifically, our advanced GERBEHRT architecture almost doubled the AVPR compared to risk factor-based models. This is encouraging in two regards: on the one hand, EHRs are routinely collected anyway. Secondly, there are clinical pictures which, in contrast to CKD, are less well understood medically and for which only limited known risk factors are available that could be used for prognosis. For predictions based on EHRs, we compared GERBEHRT with the more traditional LR and LightGBM. Per patient, GERBEHRT obtains information at which age, in which quarter, from which physician’s specialty group the patient was diagnosed with a certain disease (ICD-10-GM code and diagnostic certainty) or received a certain medication (substance and ATC code). The LR and LightGBM, on the other hand, processed only which ICD-10-GM and ATC codes, and substances were coded during the observation period and which specialty group was visited, together with the patient’s age at the observation start. Although the GERBEHRT architecture allows for a much more precise representation of the real EHR data structure, it did not outperform LightGBM by a large margin. There may be many reasons for this. One possibility could be that this type of detailed information is simply not that important for our selected prediction task. However, in contradiction to this, Krishnamurthy et al. [23] observed in their work that the prediction of CKD improves noticeably when monthly aggregated data is used instead of quarterly one. We can only speculate on the explanation. One could argue that our model would have performed better with more fine-grained temporal data as well. In addition, it is possible that not all diagnoses are recorded consistently at every doctor’s visit and that a change of doctor could possibly lead to different coding behavior, even though the underlying patient’s diseases are the same. Consequently, although GERBEHRT’s input data theoretically have a higher information content, inconsistencies, incorrect, or missing codes could reduce this advantage.

The best performing model, GERBEHRT with attention head, achieves an AUROC of 87.94 and an AVPR of 11.42. Note that a low (average) precision value is expected in the case of a class imbalance which is strongly present in our setting with a three-year incidence of moderate-to-severe CKD of 1.53% (see, for example, [61] for a study with comparable low frequencies). Hypothetically, if we were to apply our model in practice, using the F_1_ cutoff, this would mean that 27% (recall) of patients who will develop moderate-to-severe CKD in the next three years would be detected. However, it would also mean that only 14% (precision) of the patients for whom GERBEHRT predicts CKD would actually develop CKD according to the information from the EHRs and the rest would be false positives. While patients identified as CKD positive can also be considered as such almost certainly (M2Q criterion), it should be noted that CKD is often underdiagnosed. For example, the REVEAL-CKD study found a prevalence of 84.3% of undiagnosed stage 3 CKD patients in Germany [26]. Consequently, it remains to be evaluated how many of our false positives would eventually be confirmed as CKD cases upon proper screening.

We showed that our proposed GERBEHRT strategy of modeling different concepts through various features worked. This was demonstrated first by visualizing that GERBEHRT learned meaningful embeddings through the pre-training phase by clustering the learned ICD-10-GM codes and using ICD chapters as a proxy to assess semantic similarity, and second by the ablation study that showed that all features included could be used by our model and increased the predictive power. This proof of concept paves the way for the integration of additional concepts with their special properties (e.g. medical procedures) in possible subsequent studies.

Our ablation study of GERBEHRT has shown that all input features used had predictive power, although their contributions vary in magnitude. In addition to ICD-10-GM codes, age has proven to be particularly relevant. This aligns with existing knowledge that the risk of CKD increases with age [54], a piece of information that our models can use. However, while age is a strong predictor, its utility is limited, as the medical consequences for patients decrease with age. Therefore, it is interesting to note that even when age is excluded as a predictor of GERBEHRT, our model still outperforms prediction models based on risk factors or aggregated EHR statistics that include age. The quarter, medical substance, and specialty group of the physician were of similar medium importance for CKD prediction. The quarter (value between 1 and 4) might be used to get a better sense of time and order of diseases and medications, since the patient’s age was only provided in a year-wise resolution. The importance of medications could be explained by missing or incomplete diagnosis codes or diagnosis affected by certain medications. The newly introduced visited group of physicians could influence the patient’s treatment or the detection of incident diseases. The diagnostic certainty turned out to be of lower relevance. One possible explanation could be that suspected diagnoses are coded significantly less frequently than confirmed diagnoses (see Table I and Table AI in the Appendix). The ATC codes had the least positive influence on prediction performance. Possible reasons could be that ATC codes were only considered at 3-digit level and/or that medical substances alone or in combination with ICD-10-GM codes could already provide similar information content.

### C. Limitations

The data used were outpatient claims from people with statutory health insurance in Germany. This means that first hospital-coded diagnoses and prescriptions were not included, and the clinical picture of severely ill patients was only partially represented. In the case of CKD, which is treated primarily in ambulatory care [62], this should not be of enormous importance, but should nevertheless be made clear and considered in possible subsequent studies. Second, the neglect of around 11% privately insured persons in Germany [45] could also have caused a bias. And third, it is important to be aware that the EHRs used are not epidemiological data but have been generated for billing purposes. For example, the temporal resolution level for coded diagnoses is limited to the quarter, diagnoses and medications cannot be unambiguously assigned to a doctor’s visit, and the data do not contain any information on whether a patient may have died.

From a model training perspective, we found that pretraining a single GERBEHRT model took almost two weeks on a single GPU (NVIDIA GeForce RTX 2080 Ti). This makes it difficult to investigate further configurations of GERBEHRT, for instance, different resolution levels of ICD-10-GM and ATC codes.

### D. Future Directions

Early detection of CKD through screening patients with specific risk factors for proteinuria has proven effective from both medical and economic perspectives [63]–[65]. Recent work suggests that home-based albuminuria screening may also be cost-effective in the general population aged 45 to 80 [66]. Our GERBEHRT model provides estimates for the individual risk of developing CKD, allowing the selection of high-risk patients for screening, tailored to achieve a specified sensitivity. Due to its superior performance, GERBEHRT would require screening fewer patients to achieve the same sensitivity as traditional methods.

In contrast to pre-training, fine-tuning is done quickly. This means that our pre-trained GERBEHRT models now offer the possibility to be easily applied to other medical tasks, such as the prediction of further diseases, total prescription costs or the number of future visits to the physician.

Additionally, although GERBEHRT has been explicitly tailored for German EHRs, it should be possible to fine-tune and apply it in other countries for which no comparable models are yet available and which have similar population structures. For example, most countries use the ICD-10 of WHO for coding diseases, in either original or modified form, which means that a large proportion of ICD-10-GM codes should be transferable, and special properties that only apply to Germany could be modified (e.g., the diagnostic certainty could be filled with the placeholder “confirmed”).

In recent years, LLMs have been able to achieve ever better performances [67]. While these models were initially reserved for companies with enormous computing resources, smaller and yet more powerful models have also been developed and made available to the public (e.g. Bloom [68], LLama 3 [69] or GPT-NeoX [70]), and methods such as Low-Rank Adaption [71] have made it possible to fine-tune these models with limited resources. Recently, LLMs, which were fine-tuned on EHRs, were compared with Med-BERT and achieved an increase in performance for the prediction of CKD in the next medical visit or as a subsequent diagnosis [27]. This emphasizes that even the most up-to-date language models, despite billions of parameters, could be further adapted and evaluated for EHRs and that the latest developments in different research areas such as NLP should always be questioned as to whether they can be transferred to EHRs.

## V. Conclusion

In this study, we predicted incident moderate-to-severe CKD in German adults using outpatient claims data from millions of statutorily insured individuals. We developed GERBEHRT, a transformer model based on BEHRT and specifically tailored to these data, which demonstrated an improved risk assessment for CKD compared to traditional models or predictions based on individual risk factors. By providing individual CKD risk scores, GERBEHRT could support more informed decision-making by patients and doctors and could be part of targeted screening programs, reducing the number of people who must be screened to reach a specified sensitivity. Despite the improved predictive power of GERBEHRT, challenges remain in achieving highly precise individualized CKD risk estimates, and the current performance levels do not permit its broad deployment in healthcare, allowing only for targeted applications in close consultation with treating physicians. Given the high prevalence of CKD and the additional burden of an aging society in Germany, the search for solutions is becoming increasingly urgent and should be pursued further.

## Data Availability

This study is based on German Statutory Health Insurance (SHI) claims data for ambulatory care, specifically drug prescription data from pharmacies and other institutions according to Section 300 (2) of the German Social Code Book V, as well as diagnosis data of SHI-accredited physicians or psychotherapists according to Section 295 (2) of the German Social Code Book V. These data are subject to strict legal and data protection restrictions and therefore cannot be made publicly available. Access to SHI claims data is regulated by German law and requires approval from the relevant authorities. The authors are available to discuss such possibilities within the limits of research regulations.

## Appendix

**TABLE AI.** Characteristics of patients used for moderate to severe CKD prediction (8.279.440 patients in total, 901.972 patients in training and 3.688.734 in validation and test data, respectively), separated by positive and negative CKD status. Per patient, we computed properties of different features from the patient’s EHR in the observation period (2018 to 2020, with n=number/count) and built the mean, median and standard deviation (SD) over all patients with the same CKD status.

| Feature | Property | no CKD<br>(n=7.715.708)<br>Mean (SD) | CKD<br>(n=563.732)<br>Mean (SD) |
| --- | --- | --- | --- |
| Age | At first medical event | 55.6 (14.9) | 71.4 (10.9) |
| Quarters | n(with medical events) | 9.0 (3.6) | 11.3 (1.9) |
| Diagnoses | n(total) | 92.7 (94.7) | 190.6 (132.3) |
|  | n(confirmed) | 90.4 (93.3) | 187.7 (130.8) |
|  | n(suspected) | 2.3 (3.7) | 2.9 (4.5) |
|  | n(distinct ICD<sup>a</sup>) | 26.9 (19.3) | 39.6 (22.4) |
| Drugs | n(total) | 21.2 (24.6) | 52.4 (33.5) |
|  | n(distinct ATC<sup>b</sup>) | 5.0 (3.9) | 8.8 (4.3) |
|  | n(distinct substances) | 6.5 (5.7) | 11.8 (6.8) |
| Physicians | n(distinct groups) | 4.4 (2.4) | 5.2 (2.3) |

<sup>a</sup> German Modification of the 10th Revision of the International Classification of Diseases, version 2021 [44]
<sup>b</sup> Anatomical Therapeutic Chemical Classification System [46]

**TABLE AII.**
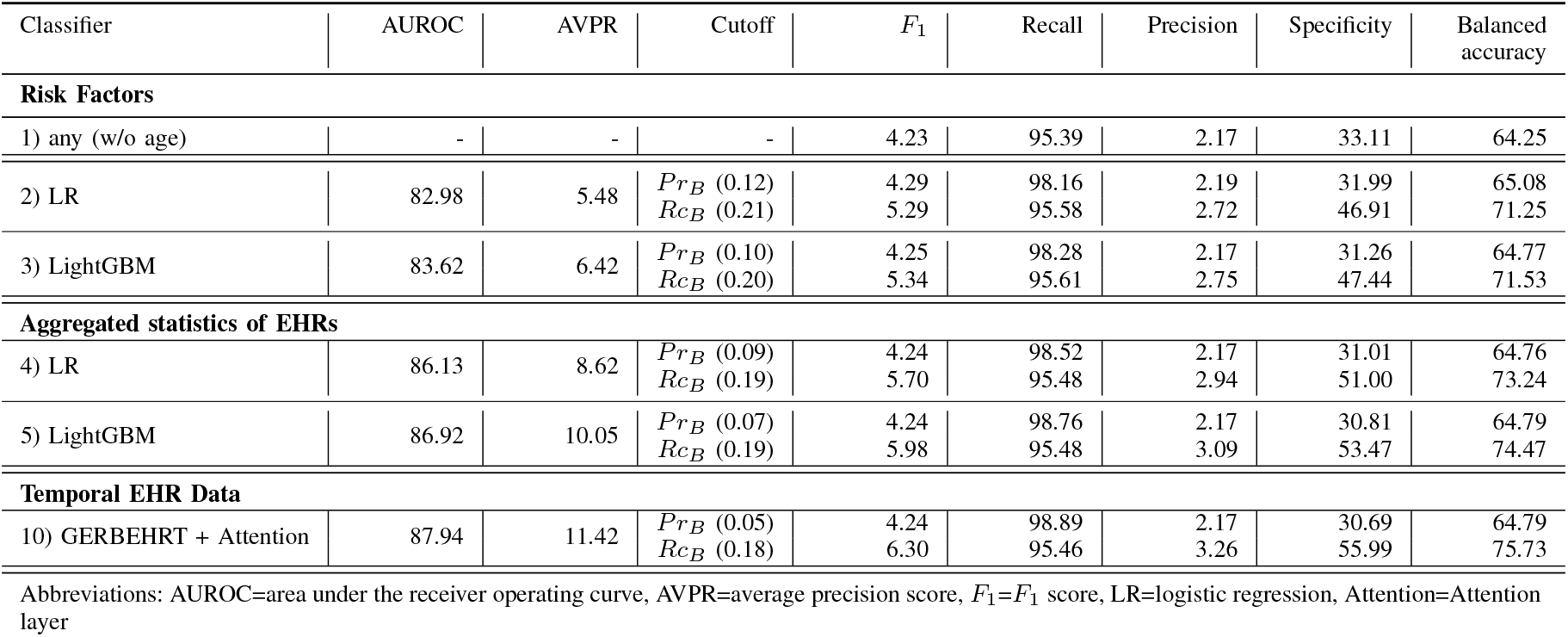
Comparison of the binary “any-risk-factor” approach with the probability returning models for moderate to severe CKD prediction on the test data set. For this, two additional prediction cutoffs were determined for the probability returning models: one that achieves at least the recall (***RcB***) and one that achieves at least the precision (***P rB***) as the baseline “any-risk-factor”-approach on the validation data set.

| Classifier | AUROC | AVPR | Cutoff | $F_1$ | Recall | Precision | Specificity | Balanced accuracy |
| --- | --- | --- | --- | --- | --- | --- | --- | --- |
| Risk Factors |  |  |  |  |  |  |  |  |
| 1) any (w/o age) | - | - | - | 4.23 | 95.39 | 2.17 | 33.11 | 64.25 |
| 2) LR | 82.98 | 5.48 | $Pr_B$ (0.12) | 4.29 | 98.16 | 2.19 | 31.99 | 65.08 |
| | | | $Rc_B$ (0.21) | 5.29 | 95.58 | 2.72 | 46.91 | 71.25 |
| 3) LightGBM | 83.62 | 6.42 | $Pr_B$ (0.10) | 4.25 | 98.28 | 2.17 | 31.26 | 64.77 |
| | | | $Rc_B$ (0.20) | 5.34 | 95.61 | 2.75 | 47.44 | 71.53 |
| Aggregated statistics of EHRs |  |  |  |  |  |  |  |  |
| 4) LR | 86.13 | 8.62 | $Pr_B$ (0.09) | 4.24 | 98.52 | 2.17 | 31.01 | 64.76 |
| | | | $Rc_B$ (0.19) | 5.70 | 95.48 | 2.94 | 51.00 | 73.24 |
| 5) LightGBM | 86.92 | 10.05 | $Pr_B$ (0.07) | 4.24 | 98.76 | 2.17 | 30.81 | 64.79 |
| | | | $Rc_B$ (0.19) | 5.98 | 95.48 | 3.09 | 53.47 | 74.47 |
| Temporal EHR Data |  |  |  |  |  |  |  |  |
| 10) GERBEHRT + Attention | 87.94 | 11.42 | $Pr_B$ (0.05)<br>$Rc_B$ (0.18) | 4.24<br>6.30 | 98.89<br>95.46 | 2.17<br>3.26 | 30.69<br>55.99 | 64.79<br>75.73 |

Abbreviations: AUROC=area under the receiver operating curve, AVPR=average precision score, $F_1=F_1$ score, LR=logistic regression, Attention=Attention layer

**TABLE AIII.** Confusion matrix counts for moderate to severe CKD prediction based on established risk factors, EHR statistics aggregated over the observation period and temporal EHRS with different (machine learning) algorithms on the test data set (CKD-positives: 56.373, CKD-negatives: 3.632.361). The highest values of AUROC and AVPR are highlighted in bold.

| Classifier | AUROC | AVPR | Cutoff | TP | TN | FP | FN |
| --- | --- | --- | --- | --- | --- | --- | --- |
| Risk Factors |  |  |  |  |  |  |  |
| 1) any (w/o age) | - | - | - | 53.777 | 1.202.557 | 2.429.804 | 2.596 |
| 2) LR | 82.98 | 5.48 | $F_1$ (0.77)<br>$J$ (0.48) | 22.180<br>46.852 | 3.305.666<br>2.559.118 | 326.695<br>1.073.243 | 34.193<br>9.521 |
| 3) LightGBM | 83.62 | 6.42 | $F_1$ (0.81)<br>$J$ (0.50) | 16.867<br>46.757 | 3.430.553<br>2.577.874 | 201.808<br>1.054.487 | 39.506<br>9.616 |
| Aggregated statistics of EHRs |  |  |  |  |  |  |  |
| 4) LR | 86.13 | 8.62 | $F_1$ (0.87)<br>$J$ (0.47) | 16.403<br>46.784 | 3.500.593<br>2.725.343 | 131.768<br>907.018 | 39.970<br>9.589 |
| 5) LightGBM | 86.92 | 10.05 | $F_1$ (0.86)<br>$J$ (0.51) | 16.281<br>46.900 | 3.515.991<br>2.760.559 | 116.370<br>871.802 | 40.092<br>9.473 |
| Temporal EHR Data |  |  |  |  |  |  |  |
| 10) GERBEHRT + Attention | 87.94 | 11.42 | $F_1$ (0.87)<br>$J$ (0.50) | 15.245<br>47.648 | 3.540.723<br>2.788.295 | 91.638<br>844.066 | 41.128<br>8.725 |

Abbreviations: AUROC=area under the receiver operating curve, AVPR=average precision score, TP=true positives, FP=false positives, TN=true negatives, FN=false negatives, $J$ =Youden’s index, $F_1=F_1$ score, LR=logistic regression, Attention=Attention layer

## References

[1] A. Levey, R. Atkins, J. Coresh, E. Cohen, A. Collins, K.-U. Eckardt, M. Nahas, B. Jaber, M. Jadoul, A. Levin et al., “Chronic kidney disease as a global public health problem: approaches and initiatives–a position statement from Kidney Disease Improving Global Outcomes,” Kidney international, vol. 72, no. 3, pp. 247–259, 2007.

[2] A. Levin, P. E. Stevens, R. W. Bilous, J. Coresh, A. L. De Francisco, P. E. De Jong, K. E. Griffith, B. R. Hemmelgarn, K. Iseki, E. J. Lamb et al., “Kidney Disease: Improving Global Outcomes (KDIGO) CKD Work Group. KDIGO 2012 clinical practice guideline for the evaluation and management of chronic kidney disease,” Kidney international supplements, vol. 3, no. 1, pp. 1–150, 2013.

[3] B. Bikbov, C. A. Purcell, A. S. Levey, M. Smith, A. Abdoli, M. Abebe, O. M. Adebayo, M. Afarideh, S. K. Agarwal, M. Agudelo-Botero et al., “Global, regional, and national burden of chronic kidney disease, 1990–2017: a systematic analysis for the Global Burden of Disease Study 2017,” The lancet, vol. 395, no. 10225, pp. 709–733, 2020.

[4] M. Girndt, P. Trocchi, C. Scheidt-Nave, S. Markau, and A. Stang, “Prävalenz der eingeschränkten Nierenfunktion,” Deutsches ärzteblatt, 2016.

[5] A. Gandjour, W. Armsen, W. Wehmeyer, J. Multmeier, and U. Tschulena, “Costs of patients with chronic kidney disease in Germany,” PloS one, vol. 15, no. 4, p. e0231375, 2020.

[6] S. Curtis and P. Komenda, “Screening for chronic kidney disease: moving toward more sustainable health care,” Current opinion in nephrology and hypertension, vol. 29, no. 3, pp. 333–338, 2020.

[7] V. A. Luyckx, K. R. Tuttle, G. Garcia-Garcia, M. B. Gharbi, H. J. Heerspink, D. W. Johnson, Z.-H. Liu, Z. A. Massy, O. Moe, R. G. Nelson et al., “Reducing major risk factors for chronic kidney disease,” Kidney international supplements, vol. 7, no. 2, pp. 71–87, 2017.

[8] P. E. Stevens, S. B. Ahmed, J. J. Carrero, B. Foster, A. Francis, R. K. Hall, W. G. Herrington, G. Hill, L. A. Inker, R. Kazancıoğlu et al., “KDIGO 2024 clinical practice guideline for the evaluation and Management of Chronic Kidney Disease,” Kidney international, vol. 105, no. 4, pp. S117–S314, 2024.

[9] N. Bertram, F. Püschner, A. S. O. Gonçalves, S. Binder, and V. E. Amelung, “Einführung einer elektronischen Patientenakte in Deutschland vor dem Hintergrund der internationalen Erfahrungen,” Berlin, Heidelberg: Springer Berlin Heidelberg, pp. 3–16, 2019.

[10] E. Rau, T. Tischendorf, and B. Mitzscherlich, “Implementation of the electronic health record in the german healthcare system: an assessment of the current status and future development perspectives considering the potentials of health data utilisation by representatives of different stakeholder groups,” Frontiers in Health Services, vol. 4, p. 1370759, 2024.

[11] H. Bang, S. Vupputuri, D. A. Shoham, P. J. Klemmer, R. J. Falk, M. Mazumdar, D. Gipson, R. E. Colindres, and A. V. Kshirsagar, “SCreening for Occult REnal Disease (SCORED): a simple prediction model for chronic kidney disease,” Archives of internal medicine, vol. 167, no. 4, pp. 374–381, 2007.

[12] G. Collins and D. Altman, “Predicting the risk of chronic kidney disease in the UK: an evaluation of QKidney® scores using a primary care database,” British Journal of General Practice, vol. 62, no. 597, pp. e243–e250, 2012.

[13] N. A. Almansour, H. F. Syed, N. R. Khayat, R. K. Altheeb, R. E. Juri, J. Alhiyafi, S. Alrashed, and S. O. Olatunji, “Neural network and support vector machine for the prediction of chronic kidney disease: A comparative study,” Computers in biology and medicine, vol. 109, pp. 101–111, 2019.

[14] J. Qin, L. Chen, Y. Liu, C. Liu, C. Feng, and B. Chen, “A machine learning methodology for diagnosing chronic kidney disease,” IEEE access, vol. 8, pp. 20 991–21 002, 2019.

[15] M. A. Islam, M. Z. H. Majumder, and M. A. Hussein, “Chronic kidney disease prediction based on machine learning algorithms,” Journal of pathology informatics, vol. 14, p. 100189, 2023.

[16] A. Perotte, R. Ranganath, J. S. Hirsch, D. Blei, and N. Elhadad, “Risk prediction for chronic kidney disease progression using heterogeneous electronic health record data and time series analysis,” Journal of the American Medical Informatics Association, vol. 22, no. 4, pp. 872–880, 2015.

[17] R. G. Nelson, M. E. Grams, S. H. Ballew, Y. Sang, F. Azizi, S. J. Chadban, L. Chaker, S. C. Dunning, C. Fox, Y. Hirakawa et al., “Development of risk prediction equations for incident chronic kidney disease,” Jama, vol. 322, no. 21, pp. 2104–2114, 2019.

[18] X. Song, L. R. Waitman, Y. Hu, A. S. Yu, D. Robins, and M. Liu, “Robust clinical marker identification for diabetic kidney disease with ensemble feature selection,” Journal of the American Medical Informatics Association, vol. 26, no. 3, pp. 242–253, 2019.

[19] C.-C. Shih, C.-J. Lu, G.-D. Chen, and C.-C. Chang, “Risk prediction for early chronic kidney disease: results from an adult health examination program of 19,270 individuals,” International Journal of Environmental Research and Public Health, vol. 17, no. 14, p. 4973, 2020.

[20] D. A. Debal and T. M. Sitote, “Chronic kidney disease prediction using machine learning techniques,” Journal of Big Data, vol. 9, no. 1, p. 109, 2022.

[21] Y. Zhu, D. Bi, M. Saunders, and Y. Ji, “Prediction of chronic kidney disease progression using recurrent neural network and electronic health records,” Scientific Reports, vol. 13, no. 1, p. 22091, 2023.

[22] G.R. Vásquez-Morales, S. M. Martinez-Monterrubio, P. Moreno-Ger, and J. A. Recio-Garcia, “Explainable prediction of chronic renal disease in the colombian population using neural networks and case-based reasoning,” Ieee Access, vol. 7, pp. 152 900–152 910, 2019.

[23] S. Krishnamurthy, K. Ks, E. Dovgan, M. Luštrek, B. Gradišek Piletič, K. Srinivasan, Y.-C. Li, A. Gradišek, and S. Syed-Abdul, “Machine learning prediction models for chronic kidney disease using national health insurance claim data in Taiwan,” Healthcare, vol. 9, no. 5, p. 546, 2021.

[24] D. Saif, A. M. Sarhan, and N. M. Elshennawy, “Deep-kidney: an effective deep learning framework for chronic kidney disease prediction,” Health Information Science and Systems, vol. 12, no. 1, p. 3, 2023.

[25] D. Saif, A. M. Sarhan, and N. M. Elshennawy, “Early prediction of chronic kidney disease based on ensemble of deep learning models and optimizers,” Journal of electrical systems and information technology, vol. 11, no. 1, p. 17, 2024.

[26] N. Tangri, T. Moriyama, M. P. Schneider, J. B. Virgitti, L. De Nicola, M. Arnold, S. Barone, E. Peach, E. Wittbrodt, H. Chen et al., “Prevalence of undiagnosed stage 3 chronic kidney disease in France, Germany, Italy, Japan and the USA: results from the multinational observational REVEAL-CKD study,” BMJ open, vol. 13, no. 5, p. e067386, 2023.

[27] O. B. Shoham and N. Rappoport, “Cpllm: Clinical prediction with large language models,” PLOS Digital Health, vol. 3, no. 12, p. e0000680, 2024.

[28] L. Rasmy, Y. Xiang, Z. Xie, C. Tao, and D. Zhi, “Med-BERT: pretrained contextualized embeddings on large-scale structured electronic health records for disease prediction,” NPJ digital medicine, vol. 4, no. 1, p. 86, 2021.

[29] J. Devlin, M.-W. Chang, K. Lee, and K. Toutanova, “Bert: Pre-training of deep bidirectional transformers for language understanding,” arXiv preprint 1810.04805, 2018.

[30] J. Shang, T. Ma, C. Xiao, and J. Sun, “Pre-training of graph augmented transformers for medication recommendation,” arXiv preprint 1906.00346, 2019.

[31] Y. Li, S. Rao, J. R. A. Solares, A. Hassaine, R. Ramakrishnan, D. Canoy,Y. Zhu, K. Rahimi, and G. Salimi-Khorshidi, “BEHRT: transformer for electronic health records,” Scientific reports, vol. 10, no. 1, p. 7155, 2020.

[32] Y. Meng, W. Speier, M. K. Ong, and C. W. Arnold, “Bidirectional representation learning from transformers using multimodal electronic health record data to predict depression,” IEEE journal of biomedical and health informatics, vol. 25, no. 8, pp. 3121–3129, 2021.

[33] P. Prakash, S. Chilukuri, N. Ranade, and S. Viswanathan, “RareBERT: transformer architecture for rare disease patient identification using administrative claims,” in Proceedings of the AAAI conference on artificial intelligence, vol. 35, 2021, pp. 453–460.

[34] C. Pang, X. Jiang, K. S. Kalluri, M. Spotnitz, R. Chen, A. Perotte, andK. Natarajan, “CEHR-BERT: Incorporating temporal information from structured EHR data to improve prediction tasks,” in Machine Learning for Health. PMLR, 2021, pp. 239–260.

[35] M. Lentzen, T. Linden, S. Veeranki, S. Madan, D. Kramer, W. Leodolter, and H. Fröhlich, “A Transformer-Based Model Trained on Large Scale Claims Data for Prediction of Severe COVID-19 Disease Progression,” IEEE Journal of Biomedical and Health Informatics, 2023.

[36] M. Rupp, O. Peter, and T. Pattipaka, “Exbehrt: Extended transformer for electronic health records,” in International Workshop on Trustworthy Machine Learning for Healthcare. Springer, 2023, pp. 73–84.

[37] Z. Yang, A. Mitra, W. Liu, D. Berlowitz, and H. Yu, “TransformEHR: transformer-based encoder-decoder generative model to enhance prediction of disease outcomes using electronic health records,” Nature communications, vol. 14, no. 1, p. 7857, 2023.

[38] E. Antikainen, J. Linnosmaa, A. Umer, N. Oksala, M. Eskola, M. van Gils, J. Hernesniemi, and M. Gabbouj, “Transformers for cardiac patient mortality risk prediction from heterogeneous electronic health records,” Scientific Reports, vol. 13, no. 1, p. 3517, 2023.

[39] M. Odgaard, K. V. Klein, S. M. Thysen, E. Jimenez-Solem, M. Sillesen, and M. Nielsen, “CORE-BEHRT: A Carefully Optimized and Rigorously Evaluated BEHRT,” arXiv preprint 2404.15201, 2024.

[40] E. C. Schneider, D. O. Sarnak, D. Squires, A. Shah, and M. M. Doty, Mirror Mirror 2017. Commonwealth Fund New York, 2017.

[41] I. Papanicolas, L. R. Woskie, and A. K. Jha, “Health care spending in the United States and other high-income countries,” Jama, vol. 319, no. 10, pp. 1024–1039, 2018.

[42] G. Ridic, S. Gleason, and O. Ridic, “Comparisons of health care systems in the United States, Germany and Canada,” Materia socio-medica, vol. 24, no. 2, p. 112, 2012.

[43] World Health Organization, International Statistical Classification of Diseases and related health problems: Alphabetical index. World Health Organization, 2004, vol. 3.

[44] Bundesinstitut für Arzneimittel und Medizinprodukte (BfArM) im Auftrag des Bundesministeriums für Gesundheit (BMG) unter Beteiligung der Arbeitsgruppe ICD des Kuratoriums für Fragen der Klassifikation im Gesundheitswesen (KKG), ICD-10-GM Version 2021, Systematisches Verzeichnis: Internationale statistische Klassifikation der Krankheiten und verwandter Gesundheitsprobleme, 10. Revision, Stand: 18. September 2020. Köln: BfArM, 2020, accessed July 17, 2025. [Online]. Available: https://www.bfarm.de/SharedDocs/Downloads/DE/Kodiersysteme/klassifikationen/icd-10-gm/vorgaenger/icd10gm2021zip.html

[45] R. Busse, M. Blümel, F. Knieps, and T. Bärnighausen, “Statutory health insurance in Germany: a health system shaped by 135 years of solidarity, self-governance, and competition,” The Lancet, vol. 390, no. 10097, pp. 882–897, 2017.

[46] World Health Organization, “Anatomical Therapeutic Chemical Classification,” accessed July 17, 2025. [Online]. Available: https://www.whocc.no/atc/structureandprinciples/

[47] T. Wolf, L. Debut, V. Sanh, J. Chaumond, C. Delangue, A. Moi, P. Cistac, T. Rault, R. Louf, M. Funtowicz et al., “Transformers: State-of-the-Art Natural Language Processing,” in Proceedings of the 2020 conference on empirical methods in natural language processing: system demonstrations, 2020, pp. 38–45.

[48] L. McInnes, J. Healy, S. Astels et al., “hdbscan: Hierarchical density based clustering.” J. Open Source Softw., vol. 2, no. 11, p. 205, 2017.

[49] A. Rosenberg and J. Hirschberg, “V-measure: A conditional entropy-based external cluster evaluation measure,” in Proceedings of the 2007 joint conference on empirical methods in natural language processing and computational natural language learning (EMNLP-CoNLL), 2007, pp. 410–420.

[50] B. L. Neuen, E. K. Yeung, J. Rangaswami, and M. Vaduganathan, “Combination therapy as a new standard of care in diabetic and non-diabetic chronic kidney disease,” Nephrology Dialysis Transplantation, vol. 40, no. Supplement 1, pp. i59–i69, 2025.

[51] S. G. Nicholls, S. M. Langan, and E. I. Benchimol, “Routinely collected data: the importance of high-quality diagnostic coding to research,” Cmaj, vol. 189, no. 33, pp. E1054–E1055, 2017.

[52] S. Drösler, J. Hasford, B.-M. Kurth, M. Schaefer, J. Wasem, and E. Wille, “Evaluationsbericht zum Jahresausgleich 2009 im Risikostrukturausgleich,” Endfassung vom, vol. 22, p. 2011, 2011.

[53] S. S. Martin, A. W. Aday, Z. I. Almarzooq, C. A. Anderson, P. Arora, C. L. Avery, C. M. Baker-Smith, B. Barone Gibbs, A. Z. Beaton, A. K. Boehme et al., “2024 heart disease and stroke statistics: a report of US and global data from the American Heart Association,” Circulation, vol. 149, no. 8, pp. e347–e913, 2024.

[54] R. Kazancioğlu, “Risk factors for chronic kidney disease: an update,” Kidney international supplements, vol. 3, no. 4, pp. 368–371, 2013.

[55] D. van Mil, L. Kieneker, I. van Geer-Postmus, G. Prins, E. Harms,N. Stoker, M. Leving, M. Oudenhuijzen, J. Kocks, R. Gansevoort et al., “1966 Participation rate and yield of screening for albuminuria in the primary care setting for early identification of chronic kidney disease (SALINE),” Nephrology Dialysis Transplantation, vol. 39, no. Supplement 1, pp. gfae069–0039, 2024.

[56] G. Ke, Q. Meng, T. Finley, T. Wang, W. Chen, W. Ma, Q. Ye, and T.-Y. Liu, “Lightgbm: A highly efficient gradient boosting decision tree,” Advances in neural information processing systems, vol. 30, 2017.

[57] W. J. Youden, “Index for rating diagnostic tests,” Cancer, vol. 3, no. 1, pp. 32–35, 1950.

[58] F. Pedregosa, G. Varoquaux, A. Gramfort, V. Michel, B. Thirion, O. Grisel, M. Blondel, P. Prettenhofer, R. Weiss, V. Dubourg et al., “Scikit-learn: Machine learning in Python,” the Journal of machine Learning research, vol. 12, pp. 2825–2830, 2011.

[59] A. P. Bradley, “The use of the area under the ROC curve in the evaluation of machine learning algorithms,” Pattern recognition, vol. 30, no. 7, pp. 1145–1159, 1997.

[60] M. Zhu, “Recall, precision and average precision,” Department of Statistics and Actuarial Science, University of Waterloo, Waterloo, vol. 2, no. 30, p. 6, 2004.

[61] V. Bali, V. Turzhitsky, J. Schelfhout, M. Paudel, E. Hulbert, J. Peterson-Brandt, J. Hertzberg, N. R. Kelly, and R. H. Patel, “Machine learning to identify chronic cough from administrative claims data,” Scientific Reports, vol. 14, no. 1, p. 2449, 2024.

[62] G. F. Weckmann, S. Stracke, A. Haase, J. Spallek, F. Ludwig, A. Angelow, J. M. Emmelkamp, M. Mahner, and J.-F. Chenot, “Diagnosis and management of non-dialysis chronic kidney disease in ambulatory care: a systematic review of clinical practice guidelines,” BMC nephrology, vol. 19, pp. 1–18, 2018.

[63] K. Borch-Johnsen, H. Wenzel, G. Viberti, and C. Mogensen, “Is screening and intervention for microalbuminuria worthwhile in patients with insulin dependent diabeteš” British Medical Journal, vol. 306, no. 6894, pp. 1722–1725, 1993.

[64] L. E. Boulware, B. G. Jaar, M. E. Tarver-Carr, F. L. Brancati, and N. R. Powe, “Screening for proteinuria in us adults: a cost-effectiveness analysis,” Jama, vol. 290, no. 23, pp. 3101–3114, 2003.

[65] T. J. Hoerger, J. S. Wittenborn, J. E. Segel, N. R. Burrows, K. Imai, P. Eggers, M. E. Pavkov, R. Jordan, S. M. Hailpern, A. C. Schoolwerth et al., “A health policy model of ckd: 2. the cost-effectiveness of microalbuminuria screening,” American journal of kidney diseases, vol. 55, no. 3, pp. 463–473, 2010.

[66] X. G. Pouwels, D. Van Mil, L. M. Kieneker, C. Boersma, R. W. van Etten, B. Evers-Roeten, H. J. Heerspink, M. H. Hemmelder, M. L. Langelaan, M. H. Thelen et al., “Cost-effectiveness of home-based screening of the general population for albuminuria to prevent progression of cardiovascular and kidney disease,” EClinicalMedicine, vol. 68, 2024.

[67] Y. Chang, X. Wang, J. Wang, Y. Wu, L. Yang, K. Zhu, H. Chen, X. Yi, C. Wang, Y. Wang et al., “A survey on evaluation of large language models,” ACM transactions on intelligent systems and technology, vol. 15, no. 3, pp. 1–45, 2024.

[68] B. Workshop, T. L. Scao, A. Fan, C. Akiki, E. Pavlick, S. Ilić, D. Hesslow, R. Castagné, A. S. Luccioni, F. Yvon et al., “Bloom: A 176b-parameter open-access multilingual language model,” arXiv preprint 2211.05100, 2022.

[69] A. Dubey, A. Jauhri, A. Pandey, A. Kadian, A. Al-Dahle, A. Letman, A. Mathur, A. Schelten, A. Yang, A. Fan et al., “The llama 3 herd of models,” arXiv preprint 2407.21783, 2024.

[70] S. Black, S. Biderman, E. Hallahan, Q. Anthony, L. Gao, L. Golding, H. He, C. Leahy, K. McDonell, J. Phang et al., “Gpt-neox-20b: An open-source autoregressive language model,” URL https://arxiv.org/abs/2204.06745, 2022.

[71] E. J. Hu, Y. Shen, P. Wallis, Z. Allen-Zhu, Y. Li, S. Wang, L. Wang, W. Chen et al., “Lora: Low-rank adaptation of large language models.” ICLR, vol. 1, no. 2, p. 3, 2022.

